# The socioeconomic gap in life expectancy in Germany: a decomposition of age- and cause-specific mortality contributions, 2003–2021

**DOI:** 10.1101/2023.12.08.23299713

**Authors:** F Tetzlaff, M Sauerberg, P Grigoriev, J Tetzlaff, M Mühlichen, J Baumert, N Michalski, A Wengler, E Nowossadeck, J Hoebel

## Abstract

**Background:** Earlier death among people in socioeconomically deprived circumstances has been found internationally and for various causes of death, resulting in a considerable life expectancy gap between socioeconomic groups. This study examines how age- and cause-specific mortality contributions to the socioeconomic gap in life expectancy have changed in Germany over time.

**Methods:** Official German population and cause-of-death statistics from 2003 to 2021 were linked to the district-level German Index of Socioeconomic Deprivation. Life-table and decomposition methods were applied to calculate life expectancy by deprivation quintile and decompose the life expectancy gap (ΔLE) between the most and least deprived quintiles into age- and cause-specific mortality contributions.

**Findings:** From 2003 to 2019, the ΔLE between the most and least deprived quintiles of districts increased from 1.1 to 1.8 years among women and from 3.0 to 3.1 years among men. Thereafter, in the COVID-19 pandemic, it increased more rapidly to 2.2 and 3.5 years respectively in 2021. The causes of death contributing most to the ΔLE were cardiovascular diseases (CVD) and cancer, with declining contributions of CVD deaths at age 70 and above and increasing contributions of cancer deaths at ages 40–74 over time. COVID-19 mortality at ages 45+ was the strongest contributor to the increase in ΔLE after 2019.

**Interpretation:** To reduce the socioeconomic gap in life expectancy, effective efforts are needed to prevent early deaths from CVD and cancer in deprived populations, with cancer prevention and control becoming an increasingly important field of action in this respect.

## Introduction

During the last century, life expectancy increased steadily in almost all high-income countries worldwide [1]. However, in the last decade, the increase in life expectancy has slowed down and mortality trends have begun to stagnate or even deteriorate (e.g. in the US and the UK) [2]. During the COVID-19 pandemic, life expectancy decreased substantially in almost all high-income countries [2-6]. Since German reunification in 1990, life expectancy (at birth) in Germany increased by 0.27 years annually until 2010, but slowed down thereafter, peaking in 2019 at 78.7 years for men and 83.5 years for women. Following the onset of the COVID-19 pandemic in 2020, life expectancy dropped substantially, reaching 78.1 years among men and 82.8 years among women in 2022 [7]. However, there are considerable regional variations in mortality levels and trends, even within German federal states [5, 8]. The reasons for these differences could be differences in economic prosperity as well as differences in population composition by socioeconomic position, exacerbated by selective migration, factors considered to be related to health-related behaviours, and health infrastructures in the regions [2, 5, 8, 9]. In general, however, the differences between the districts decreased from 1997 to 2016 due to the strong decline in mortality in the eastern parts of Germany [8].

Jasilionis et al. have suggested that, in addition to regional inequalities, widening socioeconomic inequalities in health may be the driving forces behind the current developments in life expectancy in Germany [2]. Numerous international studies have reported higher mortality among people with lower socioeconomic position (e.g. [10-12]), and the relationship between mortality and socioeconomic position has also been examined for Germany. However, as official German mortality statistics cannot be linked to individual socioeconomic information, these studies are based on survey data [11, 13] or the administrative data of the German Pension Fund (DRV) [14, 15] or health insurance data [16]. In these studies, a four-year gap in life expectancy between women with lower and higher incomes, and a nine-to-twelve-year gap between low- and high-income men was reported [11, 16]. However, these analyses have various methodological limitations, such as a small sample size or the data not being representative of all regions or population groups. At the area level, aggregated socioeconomic information is available. The highest level of regional socioeconomic deprivation is located in regions of the former German Democratic Republic [17]. Furthermore, clusters of high socioeconomic deprivation are also found in the former heavy industrial centres and structurally weak regions in western Germany (e.g. North Rhine-Westphalia, coastal regions in Lower Saxony and Schleswig-Holstein), while the lowest levels of deprivation are located in southern Germany (Baden-Wuerttemberg and Bavaria) [17].

In Germany, cardiovascular diseases and almost all common cancers (e.g. lung, colon and rectal, pancreatic, stomach, breast, prostate) are the leading causes of death [9, 18, 19]. In terms of cancer mortality, regional analyses found clear east-west differences in smoking-related and preventable-cancer mortality, with higher levels of mortality in the eastern part of Germany [9, 20]. In addition, a north-south divide was reported with lower cancer mortality in the less deprived districts in southern Germany [18]. The currently still remaining gap in life expectancy of about 1 year in 2017 between eastern and western German men was mainly due to the less favourable development of cancer and cardiovascular mortality in the eastern parts of Germany [9, 18, 21]. For women it was mainly the sharp decline in differences in cardiovascular mortality during the transformation process that led to the disappearance of differences in life expectancy between eastern and western Germany [9].

Our paper aims to shed light on the driving forces behind the socioeconomic gap in life expectancy in Germany and to provide some explanations for the current developments. To overcome the lack of information on individual socioeconomic position in the official data, we have linked the data from the official cause-of-death statistics with an area-level index of socioeconomic deprivation. Since the socioeconomic inequalities in mortality and the underlying causes of death may have been influenced by COVID-19, we also analyse trend changes during the pandemic and which age groups and causes of death contributed to it. In detail, we addressed the following questions:

- How large is the gap in life expectancy between area-level socioeconomic groups?
- Which causes of death contribute most to the gap in life expectancy between the least and most deprived districts in Germany?
- Have the area-level socioeconomic gap in life expectancy and the specific mortality contributions of different age groups and causes of death changed over recent decades?

## Methods

### Data and cause-of-death definition

For our analysis, we used the official German population numbers and cause-of-death statistics from 2003 to 2021 as provided by the Federal Statistical Office of Germany [22]. Population numbers varied between 80 and 82 million persons per year, with death figures ranging from 818,000 to 1,024,000, covering the entire German population. Causes of death are coded according to the International Statistical Classification of Diseases and Related Health Problems (10th revision). For each year and district, most prominent ill-defined causes of death identified as ICD-codes in chapter R (symptoms, signs and abnormal clinical and laboratory findings, not elsewhere classified; R00-R54, R56-R99) were proportionally redistributed among all other causes of death according to the composition of the respective diseases by sex and age group. A detailed overview of our cause-of-death grouping according to the ICD-10 coding can be found in Table S1 in the supplement.

In order to analyse socioeconomic inequalities in mortality, we linked the data from the official statistics with the area-level composite German Index of Socioeconomic Deprivation (GISD) [17] at the district level. The GISD combines the three core dimensions of the socioeconomic position: education, employment and income. Depending on their GISD score, the districts were split into quintiles. The methodology and operationalisation of the current revision [17] and codes to reproduce the GISD are publicly available (https://github.com/robert-koch-institut/German_Index_of_Socioeconomic_Deprivation_GISD). For our analyses, we used the GISD data release 2022v1.

### Statistical analysis

First, we calculated age-specific mortality rates for all-cause mortality and different groups of causes of death (Table S1, supplement). Afterwards we used standard life table techniques to calculate life expectancy [23] for both sexes and each quintile of socioeconomic deprivation. Finally, we used the stepwise decomposition approach [24] and the function *stepwise_replacement* of R package *DemoDecomp* [25] to decompose the difference in life expectancy between the most and least deprived quintiles of German districts into age- and cause-specific contributions. In addition, we calculated the deprivation-specific standardised mortality rate for the different cause-of-death groups for broader overviews of mortality developments by socioeconomic deprivation. For standardisation, we used the European Standard Population 2013 [26]. All analyses were performed using R 4.1.2. and STATA 17.

## Results

### Development of the life expectancy gap

Life expectancy increased between 2003 and 2019, with stronger gains in less deprived than more deprived districts. In the late 2010s, life expectancy tended to stagnate, especially in the most deprived quintile. This was found for both sexes, starting somewhat earlier in men than in women. The difference in life expectancy between the most and least deprived quintiles of districts (ΔLE) increased by 0.7 years for women from 2003 to 2019, resulting in a pre-pandemic gap of 1.8 years in 2019. For men, this life expectancy gap increased by 0.1 years and amounted to approximately 3.1 years in 2019 (Figure 1). In the first two years of the COVID-19 pandemic, life expectancy decreased in both more and less deprived districts, but the pace was faster in more deprived districts, resulting in a further increase in the gap of 0.4 years among both sexes after 2019 (Figure 1).

**Fig. 1.**
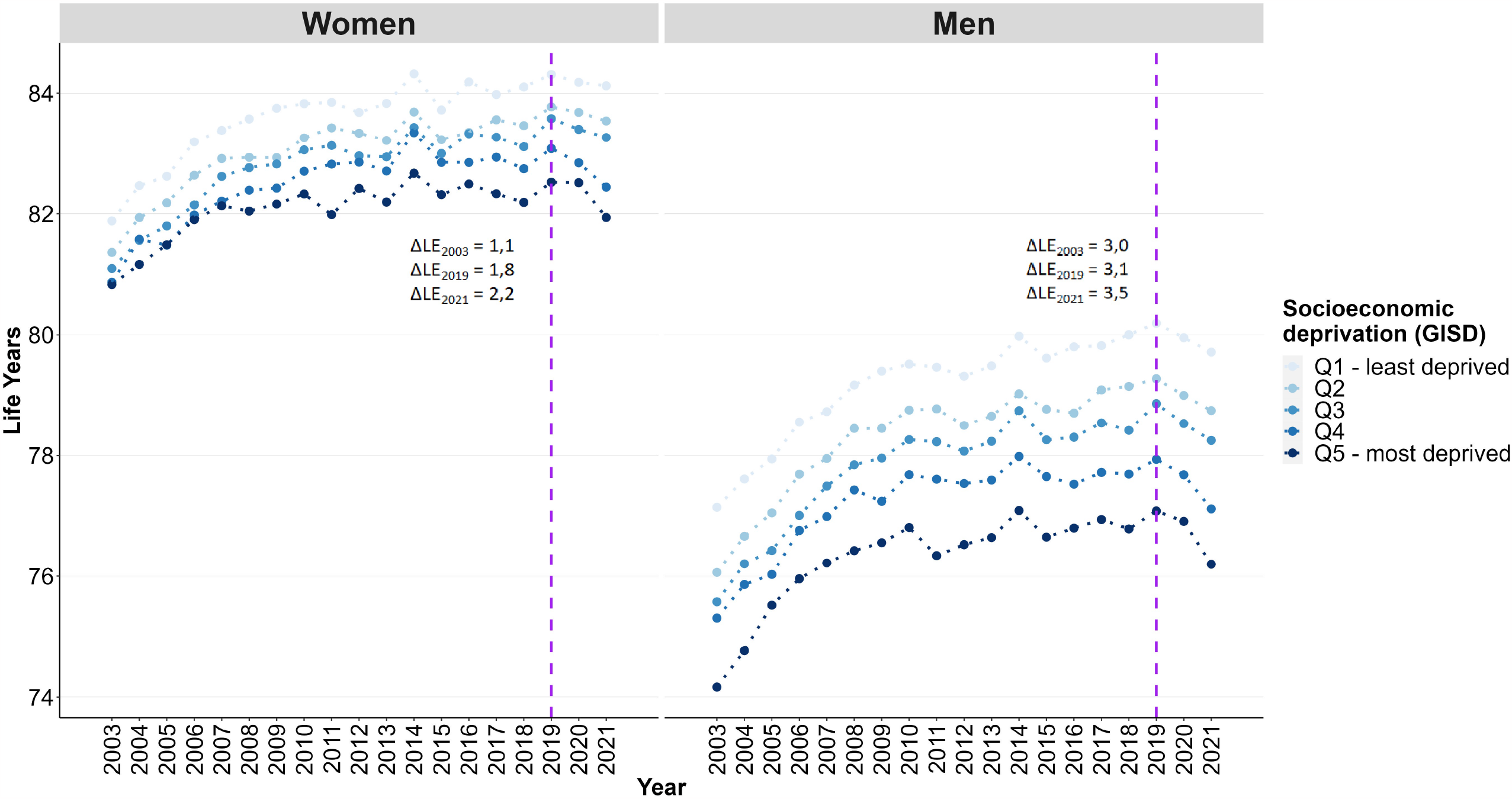
Area-level socioeconomic inequalities in life expectancy by sex and year

### Development of age- and cause-specific mortality contributions

Whereas in 2003 the life expectancy gap for women was mainly driven by mortality differences beyond the age of 60, this pattern changed in 2019, with more and more age groups below 60 contributing to the gap (Figure 2), indicating less favourable trends in premature mortality in more deprived districts. For men, the contributions shifted to higher age groups over time. This development was driven by the stronger decline in mortality inequalities at ages 0-49 (ΔLE_0-49,2003-2019_=-0.5) compared to ages 50-79 (ΔLE_50-79,2003-2019_=+0.5). The contributions of the age groups 80 and above to the life expectancy gap remained almost constant over time at 0.4 years.

**Fig. 2.**
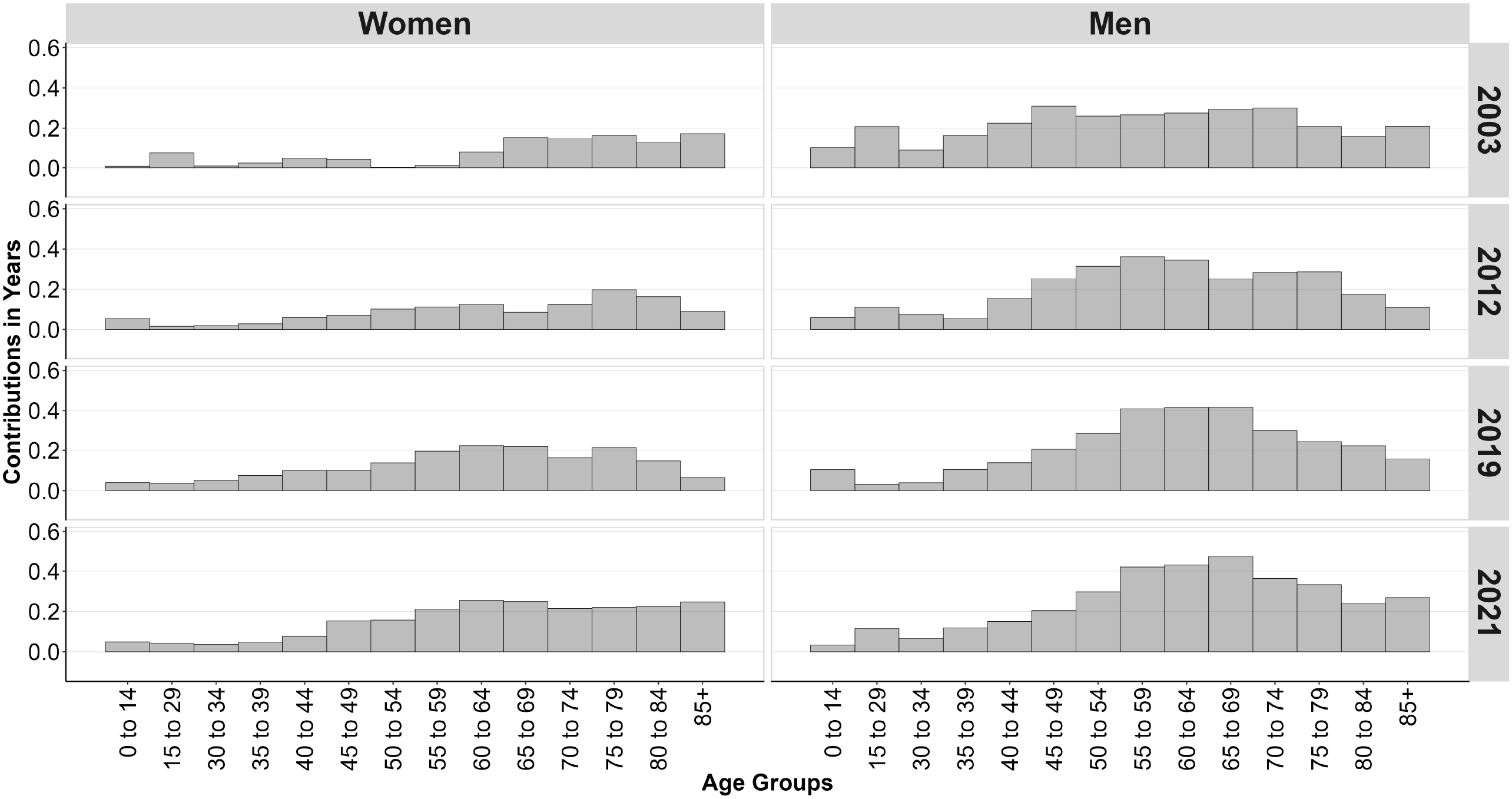
Age contributions to the gap in life expectancy between least and most deprived quintiles of districts by sex and year

In women, the increasing life expectancy gap after 2019 was mainly driven by stronger increases in mortality after age 45 in the most deprived compared to the least deprived quintile of districts (ΔLE_45+,2019-2021_=+0.4). In men, the age pattern was different: Men of a younger age (ΔLE_15-39,2019-2021_=+0.1) and above the age of 55 (ΔLE_55-85+,2019-2021_=+0.3) contributed most to the increasing life expectancy gap in 2021 (Figure 2).

Overall, reductions in mortality from cardiovascular diseases (CVD) and cancer in both sexes, and external causes of death in men, were the main drivers of gains in life expectancy between 2003 and 2019 (Figure 3 and figures S1 to S4, supplement), but not all socioeconomic groups benefited equally. For instance, we found stronger decreases in cancer mortality in the least deprived quintile, leading to increases in absolute and relative mortality inequalities. As a result, the contribution of CVD mortality to the gap in life expectancy between highest and lowest deprivation quintile decreased over time (ΔLE_CVD,women,2003-2019_=-0.3; ΔLE_CVD,men,2003-2019_=-0.4), while the socioeconomic patterning of cancer mortality widened the gap (women: ΔLE_Cancer,women,2003-2019_=+0.6; ΔLE_Cancer,men,2003-2019_=+0.4) (Figure 3). Over time, mortality from external causes decreased and inequalities almost converged in both sexes. This was especially true for the ages 0-29 years (women: ΔLE_External,women,0-29,2003-2019_=-0.03; ΔLE_External,men,0-29,2003-2019_=-0.2). At middle age, mortality from diseases of the digestive system further widened the gap in life expectancy, especially among men. At ages of 80+, mental and behavioural disorders had negative contributions to the life expectancy gap due to the increasing mortality rates from these causes in less deprived districts (Figure 3). This was mainly driven by dementia and Alzheimer’s disease, which contributed increasingly negatively to the socioeconomic life expectancy gap among these very high age groups in both sexes (Figures S5 and S6, supplement).

**Fig. 3.**
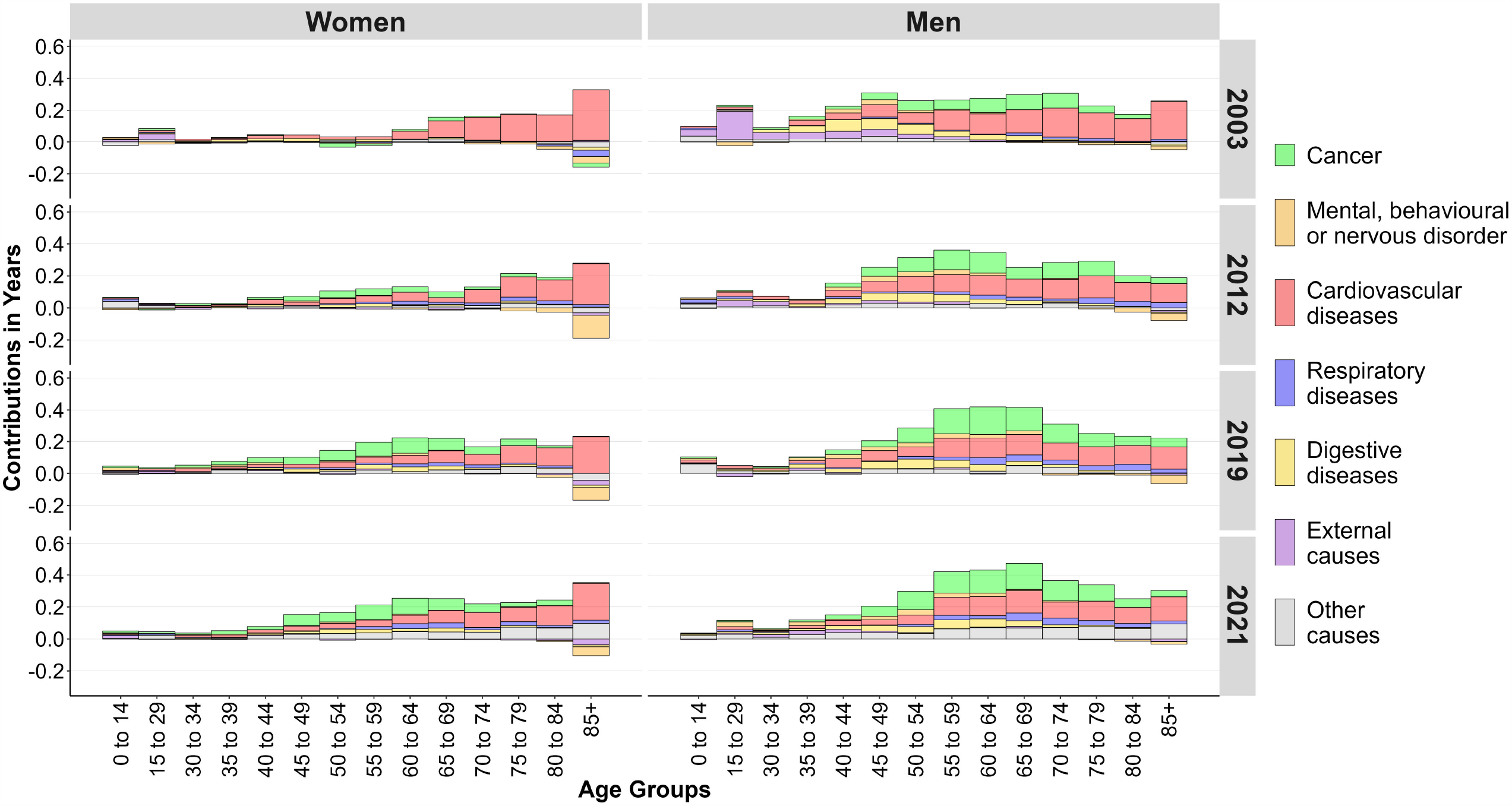
Age contributions to the gap in life expectancy between least and most deprived quintiles of districts by sex, year and cause of death

Other causes of deaths – including COVID-19 – at age 45 and above in women contributed 0.3 life years to the increasing life expectancy gap between 2019 and 2021. The remaining 0.1 life years of the increased ΔLE was mainly due to cancer, CVD and mental and behavioural disorders at age 80+ and chronic respiratory diseases at age 55+. In men, it was mainly the less favourable trend in the group of other causes of death that contributed to the increasing gap in life expectancy in the first two years of the pandemic (Figure 3).

Figure 4 shows the total contribution of the most common groups of causes of death from the ICD-10 chapters. For men, the contributions of lung cancer and preventable and treatable cancers to the life expectancy gap between the most and least deprived quintiles increased clearly up until 2019 (ΔLE_Lung Cancer,2019_=+0.4; ΔLE_Preventable Cancer,2019_=+0.2; ΔLE_Treatable Cancer,2019_=+0.2), since mortality from these cancers decreased more strongly in the least deprived districts. In women, we found increases in the contribution to the life expectancy gap only in lung and treatable cancers (ΔLE_Lung Cancer,2019_=+0.2; ΔLE_Treatable Cancer,2019_=+0.2). Ischemic heart diseases and cerebrovascular diseases narrowed the life expectancy gap over time. The contribution of ischemic heart diseases decreased to 0.5 years for men and more than halved in women to 0.3 years up until 2019, due to the stronger reduction in mortality in the most deprived districts. The contribution of cerebrovascular diseases to the life expectancy gap declined even more. It dropped by more than two-thirds for women and halved for men between 2003 and 2019 (both sexes: ΔLE_Cerebrovascular Diseases,2019_=-0.1). Among the respiratory diseases, COPD was the most important contributor to an increase in the life expectancy gap. In the first years of observation, only small inequalities in COPD mortality could be observed. Later, COPD widened the gap by 0.2 years for men and 0.1 years for women in 2019, which was due to a stronger increase in COPD mortality among women in more deprived districts and a decreasing COPD mortality among men in less deprived districts. The contribution of liver diseases, in contrast, remained almost constant over time in both sexes. The analyses also showed a reduction in the contribution of accidents to the gap among men.

**Fig. 4.**
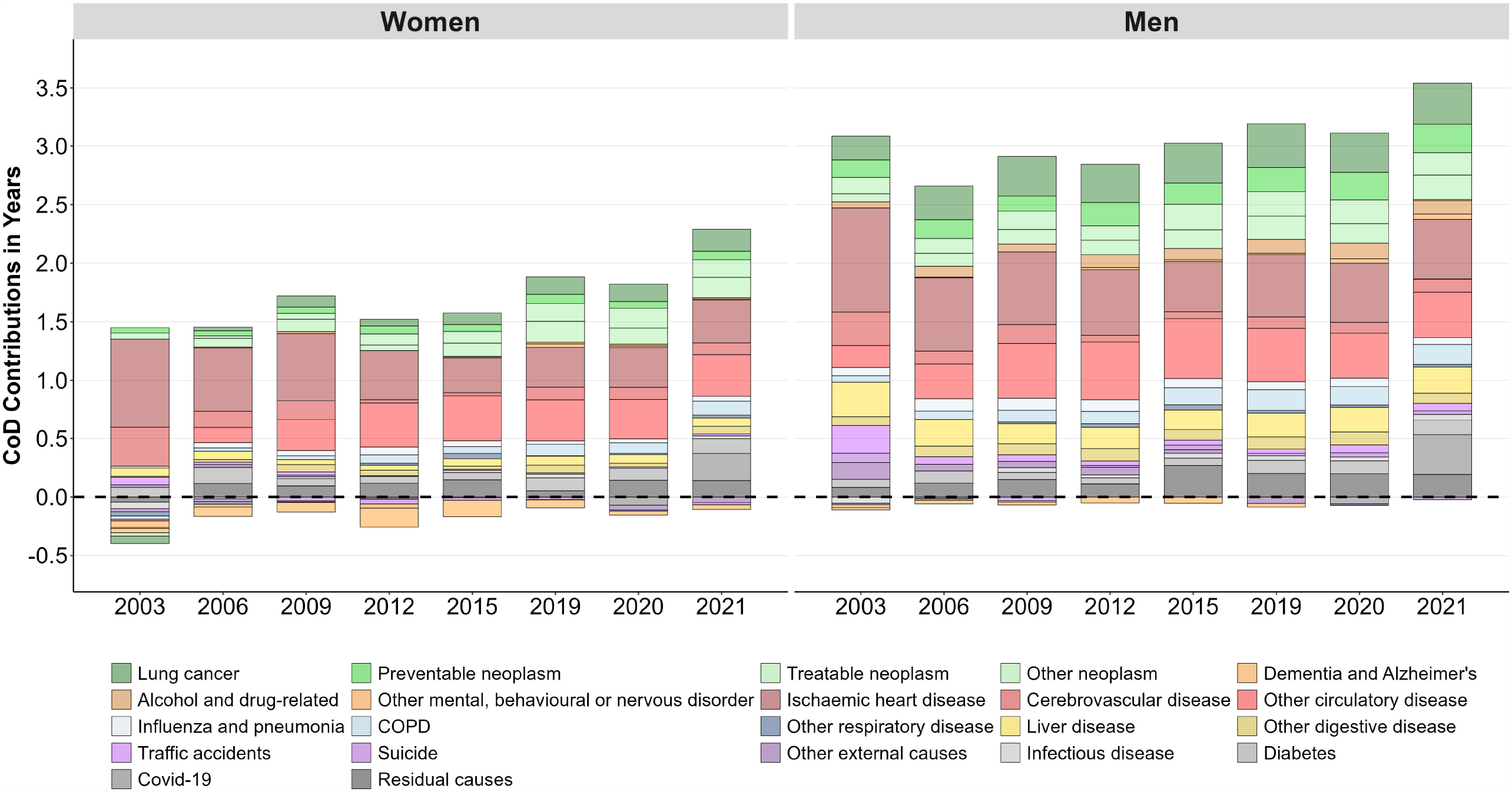
Cause-of-death contributions to the gap in life expectancy between least and most deprived quintiles of districts by sex and year

In contrast, diabetic diseases widened the gap in life expectancy slightly, resulting in a contribution from diabetic disease of approximately 0.1 years in 2019.

Overall, the trend pattern in contributing causes of deaths to the life expectancy gap observed between 2003 and 2019 continued in 2020 and 2021. However, COVID-19 as a new cause of death contributed most to the pandemic increase of the life expectancy gap between the most and least deprived quintiles of districts in both sexes (women: ΔLE_COVID-19,2021_=+0.2; men: ΔLE_COVID-19,2021_=+0.3).

## Discussion

This is the first study from Germany to examine trends of area-level socioeconomic inequalities in life expectancy and the contributions of age- and cause-specific mortality to the life expectancy gap between Germany’s least and most deprived districts. Our results show a considerable dynamic in the mortality development of individual causes of death and in their contributions to the widening life expectancy gap over time. Between 2003 and 2019, we observe an increasing gap in life expectancy between the most and least deprived districts, with an additional considerable increase in both sexes during the first years of the COVID-19 pandemic. Mortality from cardiovascular disease and cancer are the main contributors to this trend in the life expectancy gap, with a decreasing contribution from CVD deaths beyond the age of 70 and an increasing contribution of premature cancer mortality over time. After 2019, COVID-19 mortality at the age of 45 and above was one of the strongest drivers of the increase in the gap in life expectancy between the least and most deprived districts in Germany.

Earlier studies showed considerable [11] and widening socioeconomic inequalities in life expectancy in Germany [14, 16]. Our analysis goes a step further and reveals the contributions of age and cause of death to the widening gap between the most and least deprived districts in all-cause mortality in Germany between 2003 and 2021. These widening inequalities were evident in almost all considered causes of death and age groups. Compared to other high-income countries, Germany also shows a less favourable development in mortality between the ages of 50 and 79 years, but also in the age groups above. Almost half of the difference between these countries and Germany is explained by the mortality in age groups 50-79 and 80+ [2]. In our study, we were able to show that the mortality inequalities in these age groups drive socioeconomic inequalities in life expectancy and may also contribute to explaining the currently stagnating trends in life expectancy in Germany.

The sharp decline in CVD mortality is one of the most important factors behind the increase in life expectancy in recent decades [9, 21]. However, large regional and socioeconomic differences in the overall level and in the pace of declines were reported for CVD mortality [9, 17, 21]. Jasilionis et al. have suggested that the stagnation in CVD mortality and these large regional and socioeconomic inequalities in mortality, which increased over time, may be the driving forces behind the current stagnation in life expectancy [2]. Our study has shown that, although mortality from CVD is still the largest contributor to the socioeconomic gap in life expectancy, its contribution has decreased considerably over time. Cancer represents one of the most important disease groups driving the current life expectancy gap. Comparable to previous studies [18, 27, 28], we found decreasing mortality from almost all cancers. But again, not all socioeconomic groups benefitted equally from this trend, resulting in a widening socioeconomic gap in mortality. The strongest increases in inequalities were found in preventable (e.g. lung) and treatable (e.g. colon and rectal) cancers. Interpreting this against the backdrop of increased prevention efforts and nationwide cancer screenings, our analysis suggests that cancer prevention may be less effective in more deprived areas.

First results on the impact of the COVID-19 pandemic on life expectancy have reported substantial decreases in life expectancy in almost all high-income countries [2-6], which is supported by our study for Germany. In addition, our findings show that COVID-19 as a new cause of death has not only increased mortality overall, but was also accompanied by widening socioeconomic inequalities in life expectancy during the pandemic. However, after a decade in which life expectancy has increased only slightly, predictions about future trends are rather speculative, especially after the advent of COVID-19. For example, the decline in mortality from dementia, Alzheimer’s disease and chronic respiratory disease in 2020 and 2021 could be interpreted as a “harvesting effect” [29], because older age groups and people with various underlying diseases had high risks for severe COVID-19 and were hence more prone to die of COVID-19 instead of dying slightly later of dementia or other conditions.

In addition to CVD and cancer, COVID-19 emerged in 2020 and 2021 as a new main driver of the mortality gap. For each of these three disease groups, there was not only a socioeconomic mortality gradient, but also a considerably increased level of mortality from the age of 40 onwards, which had effects on both the life expectancy gap and life expectancy in Germany overall. As our findings indicate, the emergence of COVID-19 has exacerbated the expansion of area-level socioeconomic inequalities in life expectancy in Germany.

## Strength and limitations

A major limitation of the official German population and cause-of-death statistics is that they do not contain information on individual socioeconomic position. We therefore used an area-based approach, which enabled us to analyse inequalities in cause-specific mortality on a German nationwide scale. The official population and cause-of-death statistics are available in finely aggregated (district-level) as well as in detailed (subchapter-level of the ICD-10 classification) resolution. The data needed to construct the GISD are also available in excellent quality at a small-area level. However, it is important to keep in mind that we compared regional units instead of individuals, which is why we cannot rule out ecological fallacy. We analysed associations and not causalities, and associations observed at the area level may differ from associations at the individual level (atomistic fallacy).

The regional and time-related variations in the coding of causes of death could be another limitation of our study. Across regions, we see a different picture of how well causes of death are coded in Germany according to the WHO guidelines [30]. To counteract bias from miscoding, the most prominent ill-defined cause of death codes was redistributed proportionally to all valid underlying cause of death codes by age and sex for each year and district. The German cause-of-death statistics only include the underlying cause of death and do not contain any additional information on the entire causal chain or the comorbidities of the deceased. During the COVID-19 pandemic, there was a strong focus on this new cause of death, which may have led to an overestimation of COVID-19 deaths. A more detailed cause-of-death statistic could have contributed to a better understanding of coding practices and selection effects, but these are not yet available for Germany. Despite these limitations, it was precisely the methodological approach of combining area-level socioeconomic data with cause-of-death statistics that enabled us to analyse socioeconomic inequalities in life expectancy at the national scale for Germany in depth. In this approach, the GISD offers the possibility to bridge the data gap on socioeconomic inequalities in mortality in Germany.

## Conclusions

We found a clearly widening gap in life expectancy between the most and least deprived quintiles of districts in Germany. The most common contributing causes of death are CVD and cancer, with increasing contributions of cancer and decreasing contributions of CVD mortality over time. A major public-health challenge is the fact that mortality between the ages of 40 and 74 is much higher in Germany’s more deprived districts and that these inequalities represent one of the main drivers of the socioeconomic gap in life expectancy. The heterogeneity of causes of death that contribute to the socioeconomic gap in life expectancy has increased since the early 2000s, as causes other than CVD and cancer, such as COPD, diabetes and progressive neurogenerative diseases (dementia and Alzheimer’s), are becoming increasingly important. From a public-health and health-equity perspective, the kind of analysis we used in our study can help identify districts with high needs of disease prevention and control in order to reduce the socioeconomic gap in life expectancy, e.g. with equitable interventions in the field of cancer prevention and care.

## Data Availability

The official German cause-of-death data analysed in this study were provided by the Research Data Centre of the Statistical Offices. At the lowest available level of area aggregation, the data are subject to strict data protection by law and are not publicly available. However, the cause-of-death data are available upon request and application to the Research Data Centre of the Statistical Offices.
The German Index of Socioeconomic deprivation is published under CC by 4.0 licence and is freely accessible via the GitHub website of the Robert Koch-Institute.

https://github.com/robert-koch-institut/German_Index_of_Socioeconomic_Deprivation_GISD

## Declarations

### Author Contributions

FT: Conceptualisation, Data Curation, Formal Analysis, Investigation, Methodology, Project Administration, Visualization, Writing – original draft; MS: Conceptualisation, Formal Analysis, Methodology, Writing – review & editing; PG: Conceptualisation, Methodology, Writing – review & editing; JT: Conceptualisation, Methodology, Writing – review & editing; MM: Conceptualisation, Methodology, Writing – review & editing; JB: Writing – review & editing; NM: Writing – review & editing; AW: Writing – review & editing; EN: Writing – review & editing; JH: Conceptualisation, Formal Analysis, Methodology, Supervision, Writing – review & editing

### Data Availability Statement

The official German cause-of-death data analysed in this study were provided by the Research Data Centre of the Statistical Offices. At the lowest available level of area aggregation, the data are subject to strict data protection by law and are not publicly available. However, the cause-of-death data are available upon request and application to the Research Data Centre of the Statistical Offices.

The German Index of Socioeconomic deprivation is published under CC by 4.0 licence and is freely accessible via the GitHub website of the Robert Koch-Institute (https://github.com/robert-koch-institut/German_Index_of_Socioeconomic_Deprivation_GISD).

