## Supplemental Material for "The socioeconomic gap in life expectancy in Germany: a decomposition of age- and cause-specific mortality contributions, 2003–2021"

**Tab. S1 Cause-of-death Classification**

| # | Detailed category | ICD-10 | Grouped category |
|---|---|---|---|
| 1 | Infectious disease | A00-B99 | Other causes (7) |
| 2 | Lung cancer* | C33-C34 | Cancer (1) |
| 3 | Preventable neoplasm | C00-C14, C15-C16, C22, C32 | Cancer (1) |
| 4 | Treatable neoplasm** | C18-C21, C40-C41, C43-C44, C50, C53-C55, C61-C62, C67, C69, C73, C81, D10-D36 | Cancer (1) |
| 5 | Other neoplasm | Residual C codes, D00-D09, D37-D48 | Cancer (1) |
| 6 | Diabetes | E10-E14 | Other causes (7) |
| 7 | Dementia and Alzheimer's | F01, F03, G30 | Mental, behavioural or nervous disorder (2) |
| 8 | Alcohol and drug-related | F10-F19 | Mental, behavioural or nervous disorder (2) |
| 9 | Other mental, behavioural or nervous disorder | Residual F-G codes | Mental, behavioural or nervous disorder (2) |
| 10 | Ischaemic heart disease | I20-I25 | Cardiovascular diseases (3) |
| 11 | Cerebrovascular disease | I60-I69 | Cardiovascular diseases (3) |
| 12 | Other circulatory disease | Residual I codes | Cardiovascular diseases (3) |
| 13 | Influenza and pneumonia | J09-J22 | Respiratory diseases (4) |
| 14 | COPD | J41-J44 | Respiratory diseases (4) |
| 15 | Other respiratory disease | Residual J codes | Respiratory diseases (4) |
| 16 | Liver disease | K70-K77 | Digestive diseases (5) |
| 17 | Other digestive disease | K00-K69, K80-K99 | Digestive diseases (5) |
| 18 | Covid-19 | U07 | Other causes (7) |
| 19 | Traffic accidents | V01-V99 | External causes (6) |
| 20 | Suicide | X60-X84 | External causes (6) |
| 21 | Other accidents | W00-W99, X00-X59, Y35-Y98 | External causes (6) |
| 22 | Residual causes*** | Residual D-E codes; all H, L, M, N, O, P, Q codes | Other causes (7) |

* Lung cancer is also preventable but due to its significance and divergent trends among men and women, it should be studied separately.

** Sex-specific cancers (breast, uterus, cervix uteri, prostate, testis) included; leukaemia (C91-C95) not included because widely amenable only at ages below 45

*** Many infant deaths (P00-P96, Q00-Q99) included in residual group, not separately. If we should ever consider analysing trends in infant mortality, it would probably make more sense to just look at mortality below age 1 instead of selected causes.

### Fig.S1 Time trends in age-standardised mortality in men by regional socioeconomic deprivation, 2003-2021

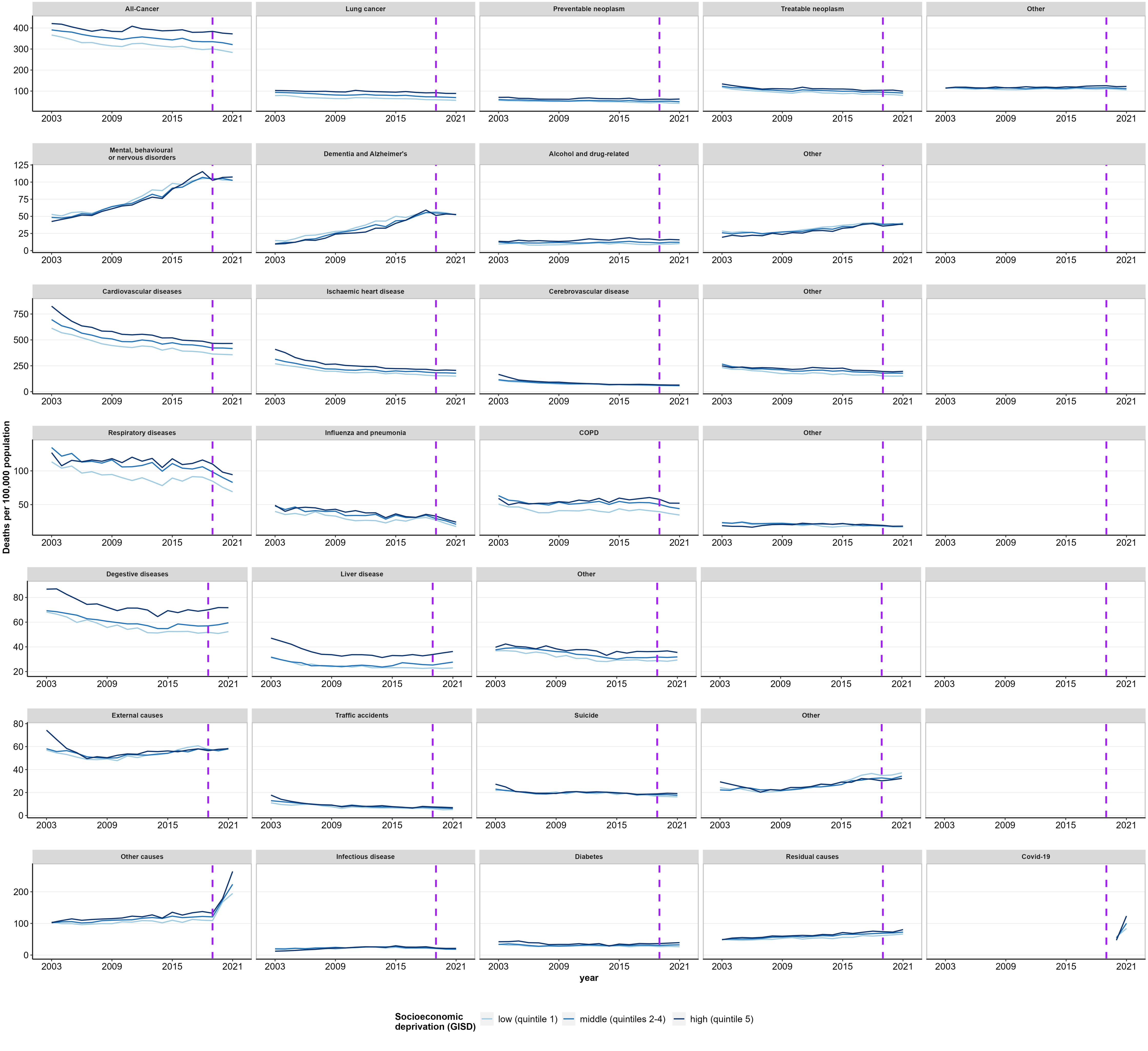

**Fig.S2 Time trends in age-standardised mortality in women by regional socioeconomic deprivation, 2003-2021**

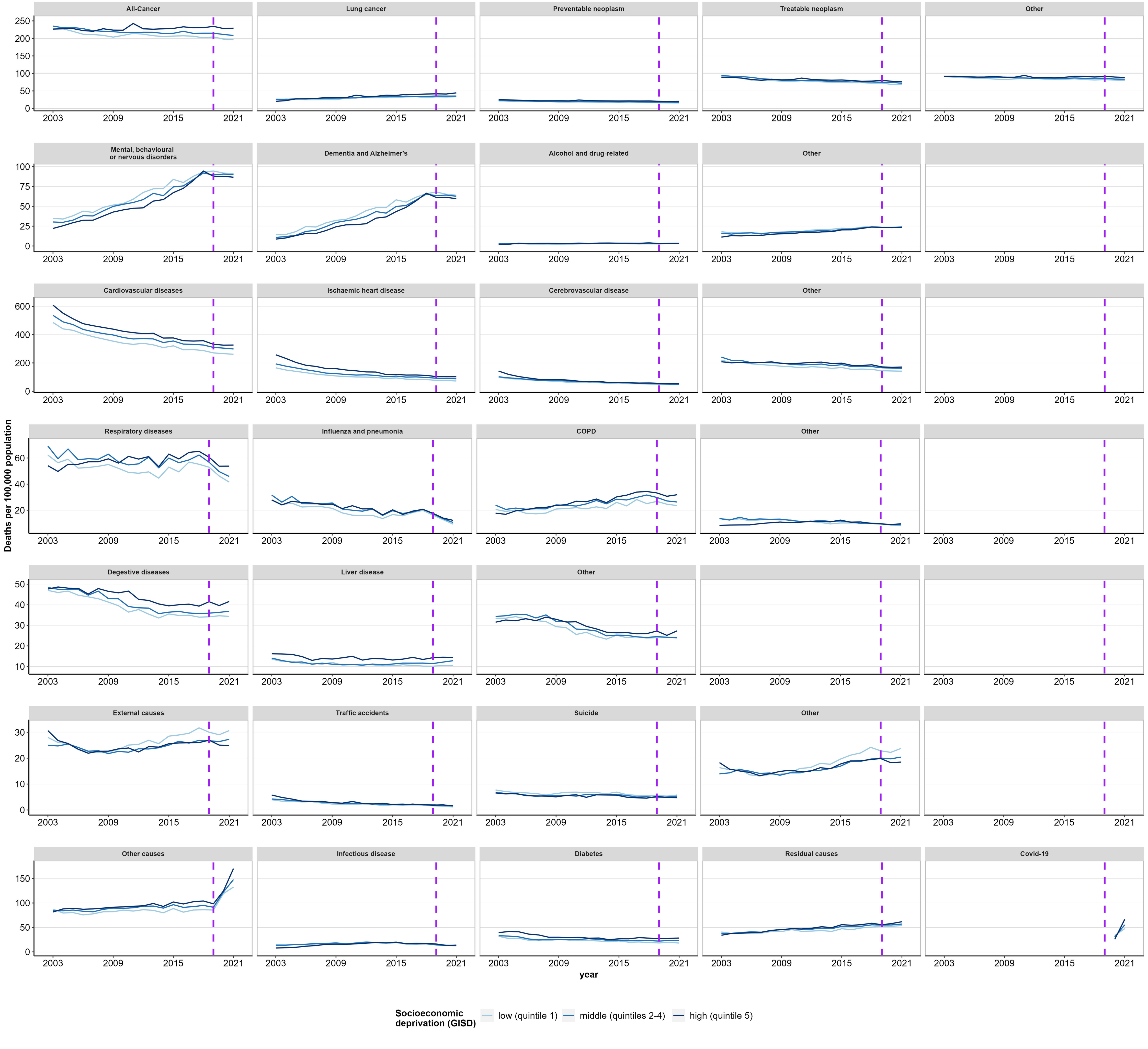

Fig. S3 Age contribution to the development of life expectancy between 2003 and 2019 in least and most deprived quintile of area-based socioeconomic deprivation by sex and cause of death

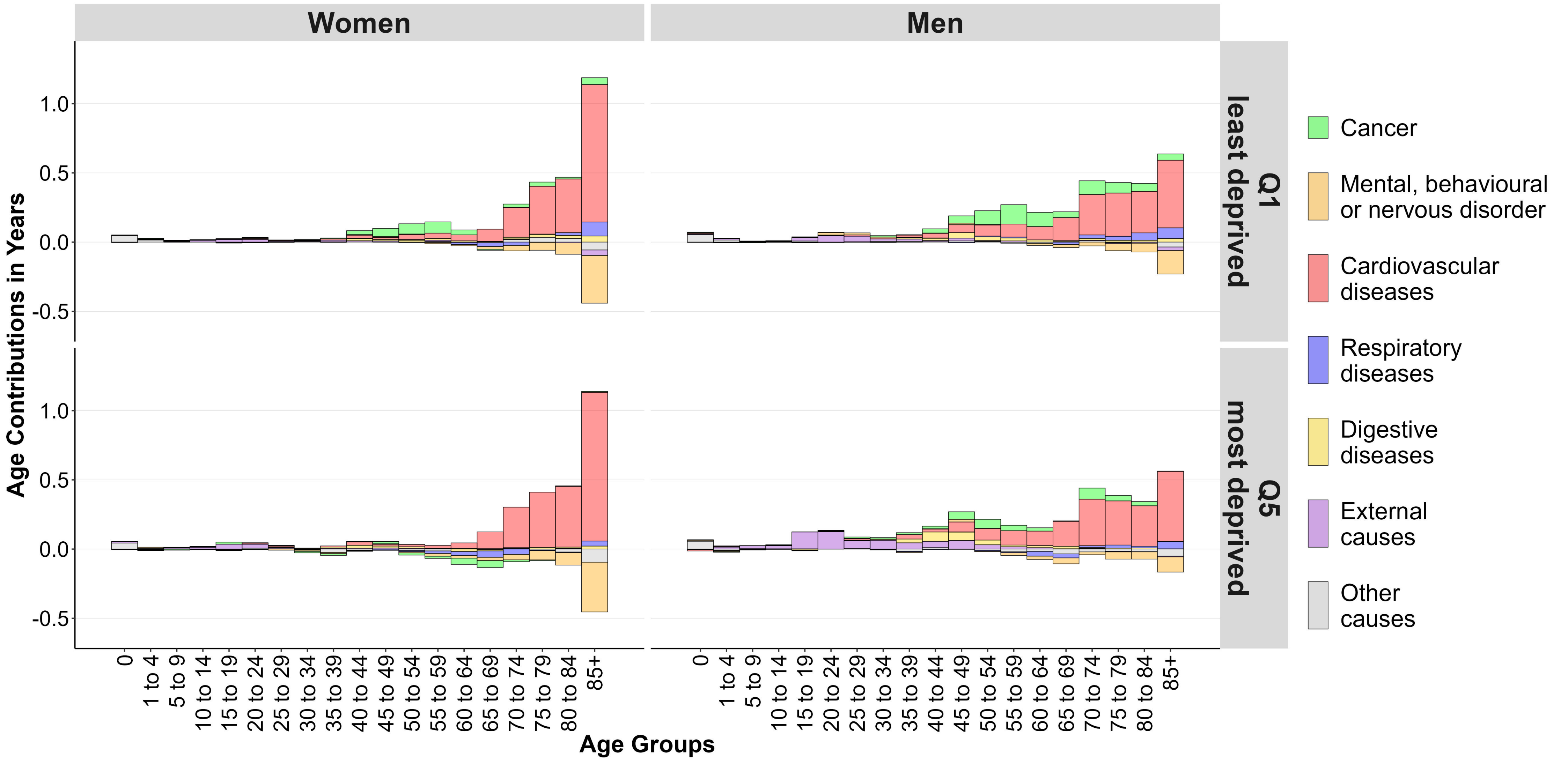

**Fig. S4 Cause of death contribution to the development of life expectancy between 2003 and 2019 in least and most deprived quintile of area-based socioeconomic deprivation by sex**

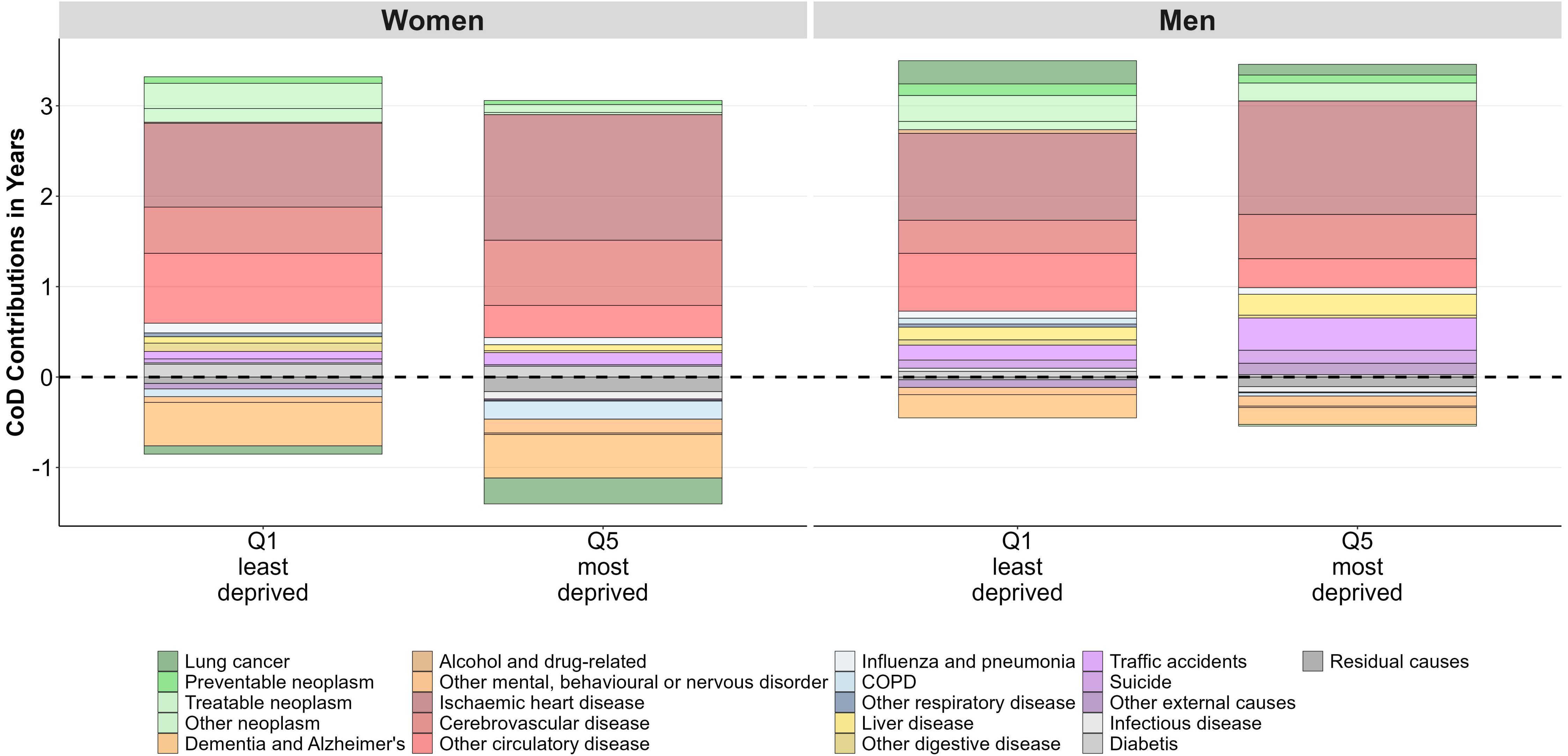

**Fig.S5 Age contributions to the gap in life expectancy in men between least and most deprived quintile of area-based socioeconomic deprivation by cause of death in 2003 and 2019**

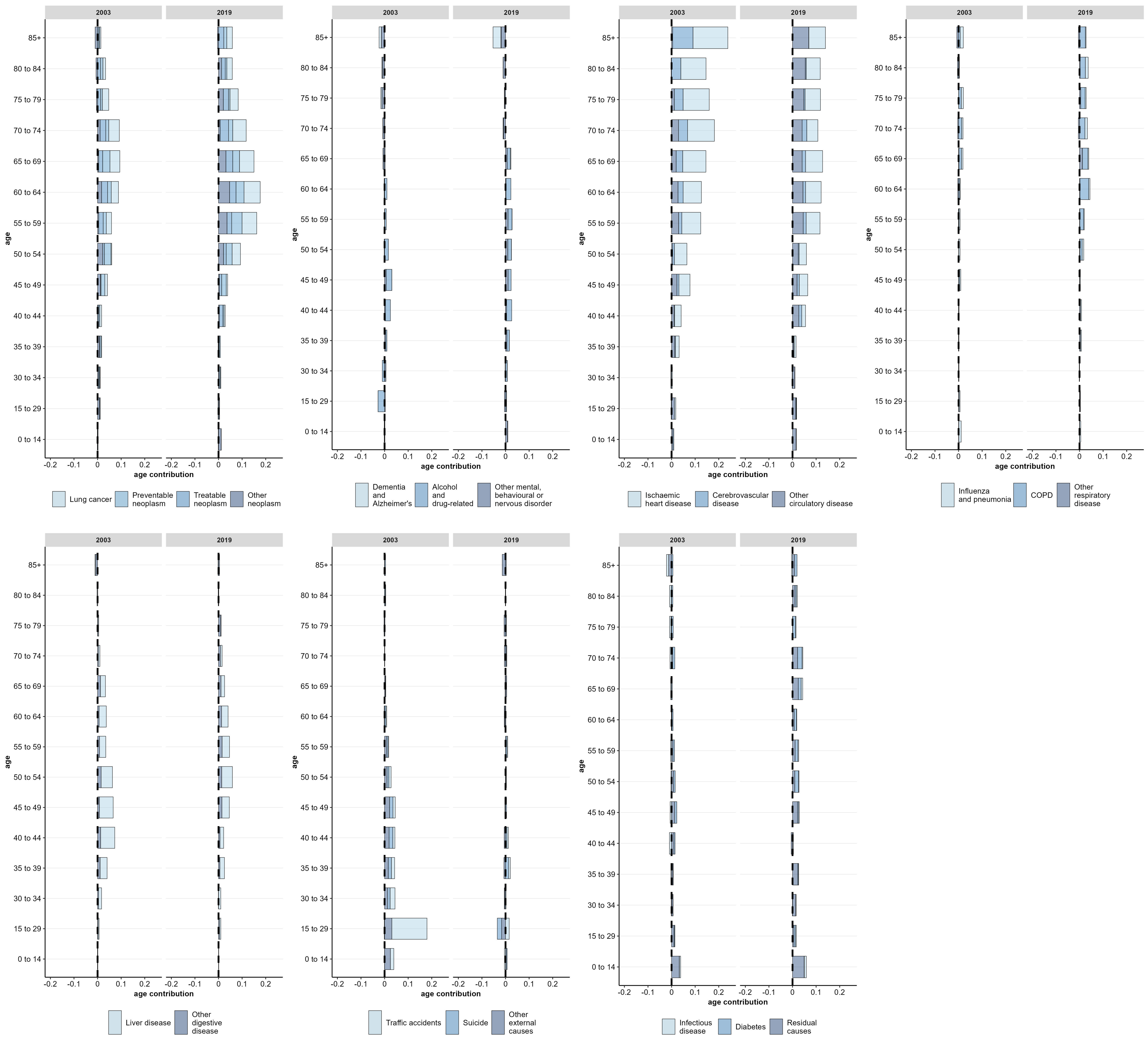

**Fig.S6 Age contributions to the gap in life expectancy in women between least and most deprived quintile of area-based socioeconomic deprivation by cause of death in 2003 and 2019**

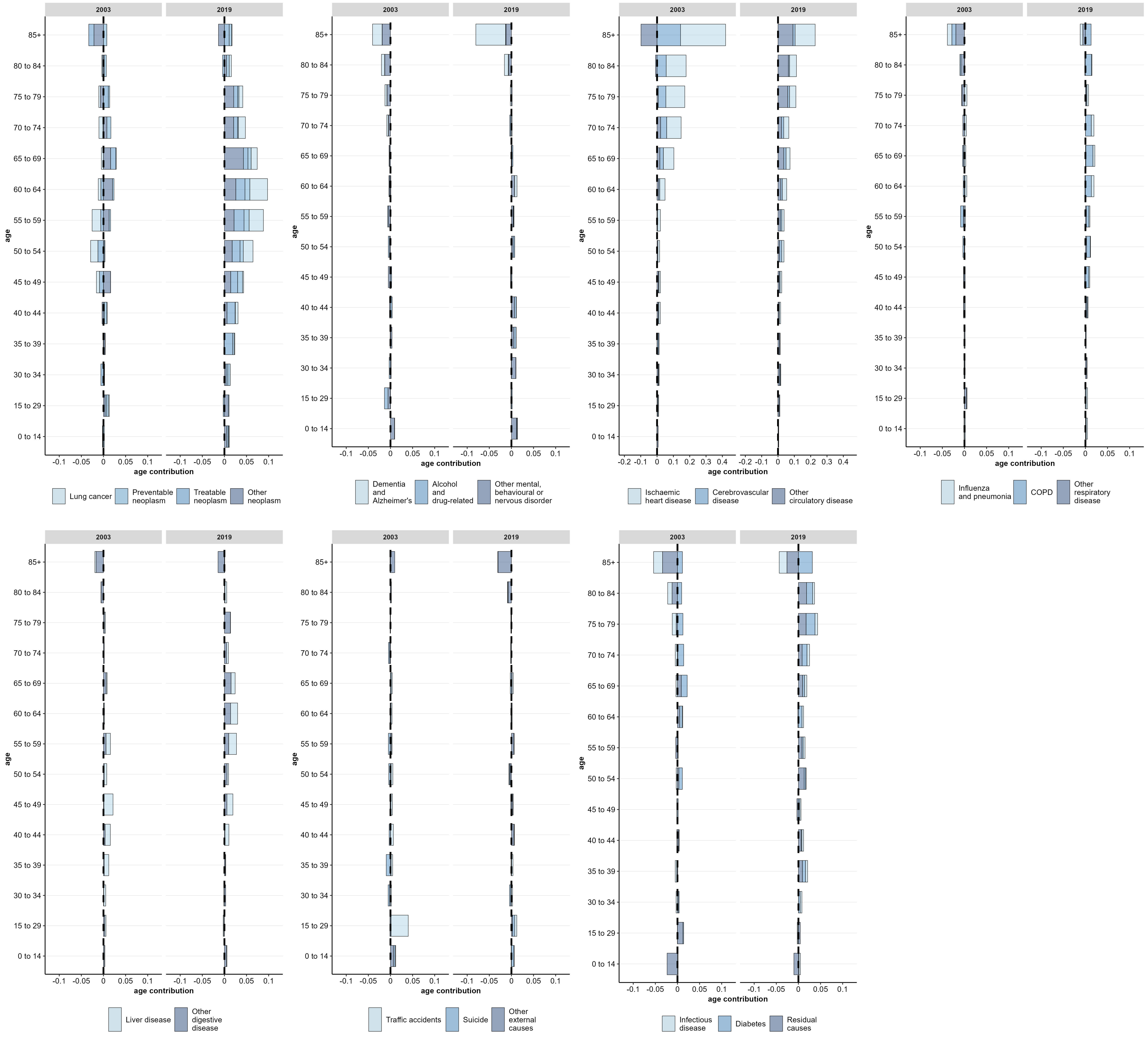
